# COVID-19 IN INDIA: MODELLING, FORECASTING AND STATE-WISE COMPARISON

**DOI:** 10.1101/2020.06.15.20131375

**Authors:** Ankitha Jose, Ashid Salim, Silpa Subhash, Noel George

## Abstract

COVID-19 has turned the whole world upside down economically and socially. COVID-19 pandemic has caused around five crores of cases and three lakhs deaths globally as of 27 May 2020. This paper adopts four mathematical growth models. Basic models are encouraged because these models can make predictions with the available data and variables in the current scenario of COVID-19 pandemic. The best-fitted model is identified in accordance with the value of the coefficient of determination. As per the best model, there might be greater than 16 lakhs cases at the infection end in India. After predicting the future size of the pandemic, we analyzed how the disease severity varies among the Indian states and union territories using Case Fatality Rates (CFR).

## I. Introduction

A new microscopic organism, familiar, yet very strange has gripped almost all the countries of the world. About five months have elapsed since it was first identified in the Wuhan province of China and yet it spreads with an unprecedented pace and an unbeatable ferocity. It has been named as severe acute respiratory syndrome coronavirus 2 (SARS-COV-2), which causes novel coronavirus disease (COVID-19) [11]. Coronaviruses are a group of related Ribonucleic Acid (RNA) viruses belonging to the family of Coronaviridae which can cause respiratory tract infections that can sometimes be lethal [10]. As of 26 May 2020, 54,04,512 cases and 3,43,514 deaths have been reported worldwide, and 4,167 deaths amongst the 1,45,380 cases reported in India only. In order to control the spread of the virus, strict sanitary measures were implemented in most countries.

In India, the first case of COVID-19 was documented on 30 January 2020. There were only 500 cases reported in India, when government ordered a 14-hour voluntary public curfew on 22 March 2020, followed by four phases of lockdown, which WHO mentioned India’s immediate decision on lockdown as “tough and timely” [7]. Along with control of the disease, accurate prediction of its spread can drastically limit its adverse effects. Thus, mathematical models have been used to predict its spread throughout the globe. In India, based on the transmission dynamics of COVID-19, mathematical models have been established to predict the size of the pandemic [4]. Case fatality rate (CFR) is also a prime concern to understand the trend of COVID-19 in Indian states and union territories.

In this study, the dynamic nature of the virus and its spread are used in mathematical growth models to estimate the expected number of cases over time in the country. Thereafter Case fatality rates among the different Indian states and union territories estimated are used to assess the varying disease severity among them.

## II. Materials And Methods

Indian COVID-19 data, consist of cumulative number of cases, was extracted from World Health Organization (WHO). The data in this analysis starts from 31 January 2020 and it ends at 10 May 2020. The prime source of the state-wise Indian data was the official website of government of India. Here we employed four mathematical growth models to estimate the number of cases that India could be facing in the future. The mathematical growth models develop a differential equation that makes use of the dynamic nature of the disease and the rate of its spread to have a better future prediction.

### Growth models

In the case of COVID-19 pandemic, **exponential growth** gives the number of cases at a particular point in time. There are several epidemiological studies suggesting that the initial stage of a pandemic follows exponential growth [5].

The mathematical model for exponential growth is:

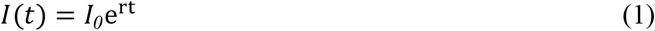

Where ***I(t)*** is the number of infections at time *t*. ***I***_***0***_ is the initial number of cases, ***r*** is the number of people infected by each sick person (growth factor), which is an important measure for the spread [9,6]

The COVID-19 outbreak can be tracked using a **logistic growth model**. It was widely utilized for modelling population growth with limited resources and space. It is commonly used for predicting the probability of occurrence of a disease based on its risk factor.

Logistic growth can be expressed as:

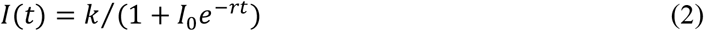

Where ***I(t)*** is the total number of infections at time t. *I*_*0*_ is the initial number of cases, ***k*** are fitting constant and ***r*** is the maximum number of cases [1].

COVID-19 growth resembles **Gompertz growth** when there is a slow growth at the beginning and end of time. This model gives the best fit when the dataset follows a smooth curve [8].

Gompertz growth is given by;

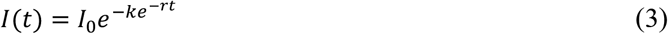

Where ***I***_***0***_ is the initial number of cases, ***k*** is the carrying capacity of an epidemic, ***r*** is the growth rate [2].

The spread law of COVID-19 pandemic and associated factors can be studied using **Bertalanffy model**. It deeply explains the factors affecting growth.

Bertalanffy growth model is;

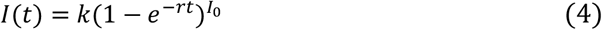

Where ***I***_***0***_ is the initial number of cases, ***k*** is the carrying capacity of an epidemic, ***r*** is the growth rate [8].

The fitting ability of growth models is compared using square of the correlation (r), the coefficient of determination (R^2^). When R^2^ is 1, the model is a good fit for the available data. A value close to or equal to 0 indicates that there is no relation between the predictor and predicted variable [8].

It is necessary to look at the spread of the disease in each state separately along with the country as a whole, given India’s size and diversity. We analyzed how disease severity varies among the selected Indian states and union territories using Case Fatality Rates (CFR).

For **Case Fatality Rate** estimation, all Indian states and union territories, which have reported at least 100 cases as per the report of 21 May 2020, are selected. Case fatality rate gives the proportion of people who die from a particular disease out of all the infected people during a certain period. CFR ranges between 0 and 1 (0% and 100%); it measures the risk of a disease in terms of disease mortality [3]. CFR=0 doesn’t imply the effect of the disease is zero, it only means that the considered disease hasn’t caused any deaths in that region to date. Quartile values are calculated using CFRs for grouping. Now, we can map out the four groups formed from these quartiles: quartile 1, quartile 2, quartile 3, and quartile 4.

Data were analyzed using python 3.8.2 (community version). The Exponential model, Logistic model, Gompertz model, and Bertalanffy model are used to model COVID-19 in India to predict the epidemic situation. Curve fitting method is adopted for the estimation of model parameters. The least square method is used for curve fitting. The error sum of squares between these observed data and predicted data can be reduced using the least squares method [8]. The best model is selected by evaluating R^2^ values.CFR values are calculated for the selected Indian states and union territories. Grouping of Indian states and union territories is done based on the quartiles calculated from the CFR values.

## III. RESULTS

Fitting analysis is done using mathematical growth models. The initial time, the time at which the first case reported is taken as zero (t_0_=0). Model parameters were estimated utilizing curve fitting.

Parameters estimated are given in table 1. Coefficients of determination values based on the fitted models are given in table 2. Here the model with the largest R^2^ value is the Bertalanffy growth model. The graphical representation of the fitted growth models is given in Figures 1, 2, 3, and 4. Table 3 represents Case fatality rates of selected states and union territories. Groups constructed on the basis of these values are given in table 4.

**Table 1:** COVID-19 prediction results in Exponential Model, Logistic Model, Gompertz Model and, Bertalanffy mode

| Model | Parameter | Value Of The Parameter |
| --- | --- | --- |
| Exponential model | $I_0$ | 0.3995 |
| | $r$ | 0.1280 |
| Logistic model | $I_0$ | 13642.0533 |
| | $r$ | 0.0949 |
| | $k$ | 124935.6670 |
| Gompertz model | $I_0$ | 20.9452 |
| | $r$ | 0.0211 |
| | $k$ | 789143.136 |
| Bertalanffy model | $I_0$ | 12.3339 |
| | $r$ | 0.0146 |
| | $k$ | 1609200.51 |

**Table 2:** Coefficient of determination (R^2^) based on fitted models.

| Growth Models | Fitted Growth Models | $R^2$ Values |
| --- | --- | --- |
| Exponential growth model | $I(t) = 0.3995 * e^{0.1280*t}$ | 0.1605 |
| Logistic growth model | $I(t) = \frac{0.0949}{1 + 13642.0533 * e^{-124935.6670*t}}$ | 0.9976 |
| Gompertz growth model | $I(t) = 789143.1360 * e^{-20.9452 * e^{-0.0211t}}$ | 0.9989 |
| Bertalanffy growth model | $I(t) = 1609200.51(1 - e^{-0.0146t})^{12.3339}$ | 0.9990 |

**Table 3:** Case fatality rates of selected states and union territories.

| State/Union Territories | Cumulative number of confirmed cases | Cumulative number of confirmed deaths | CFR |
| --- | --- | --- | --- |
| Andhra Pradesh | 2532 | 52 | 2.0537 |
| Assam | 142 | 4 | 2.8169 |
| Bihar | 1498 | 9 | 0.6008 |
| Chandigarh | 200 | 3 | 1.5 |
| Chhattisgarh | 101 | 0 | 0 |
| Delhi | 10554 | 168 | 1.5918 |
| Gujarat | 12140 | 719 | 5.9225 |
| Haryana | 964 | 14 | 1.4523 |
| Jammu And Kashmir | 1317 | 17 | 1.2908 |
| Jharkhand | 231 | 3 | 1.2987 |
| Karnataka | 1397 | 40 | 2.8633 |
| Kerala | 642 | 4 | 0.6230 |
| Madhya Pradesh | 5464 | 258 | 4.7218 |
| Maharashtra | 37136 | 1325 | 3.5679 |
| Odisha | 978 | 5 | 0.5112 |
| Punjab | 2002 | 38 | 1.8981 |
| Rajasthan | 5845 | 143 | 2.4465 |
| Tamil Nadu | 12448 | 84 | 0.6748 |
| Telangana | 1634 | 38 | 2.3256 |
| Tripura | 173 | 0 | 0 |
| Uttarakhand | 111 | 1 | 0.9009 |
| Uttar Pradesh | 4926 | 123 | 2.4969 |
| West Bengal | 63624 | 250 | 8.4431 |

**Table 4:** CFR value-based grouping of states /union territories

| Quartile 1 | Quartile 2 | Quartile 3 | Quartile 4 |
| --- | --- | --- | --- |
| CFR below or equal to Q1 (Min -0.7878) | CFR from 0.7878 and below 1.5918 | CFR from 1.5918 and below 2.6569. | CFR from 2.6569 to 8.4431 |
| Bihar | Chandigarh | Andhra Pradesh | Assam |
| Chhattisgarh | Haryana | Delhi | Gujarat |
| Kerala | Jammu And Kashmir | Punjab | Karnataka |
| Odisha | Jharkhand | Rajasthan | Madhya Pradesh |
| Tamil Nadu | Uttarakhand | Telangana | Maharashtra |
| Tripura |  | Uttar Pradesh | West Bengal |

**Figure 1:**
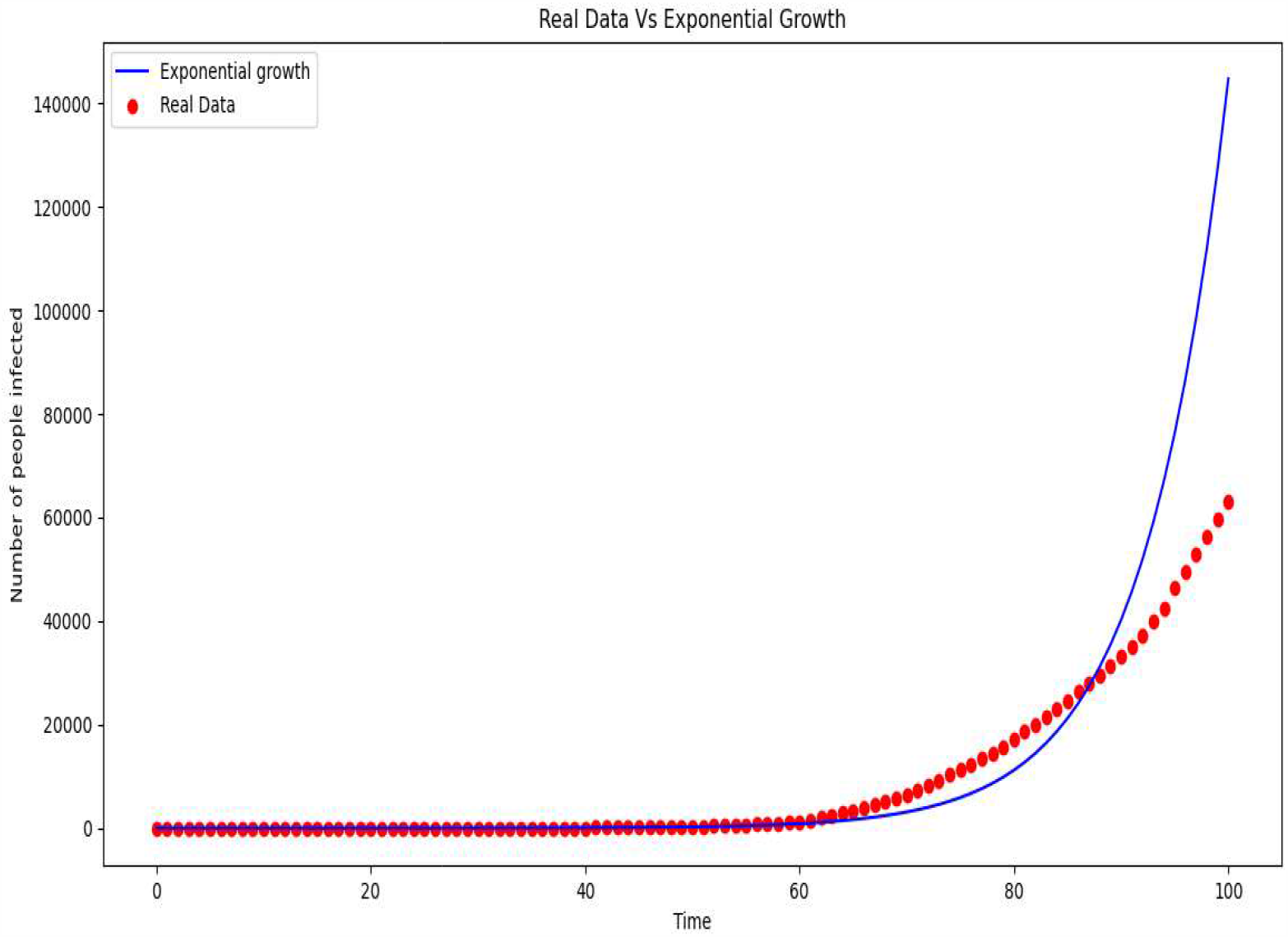
Exponential growth curve of COVID-19 in India

**Figure 2:**
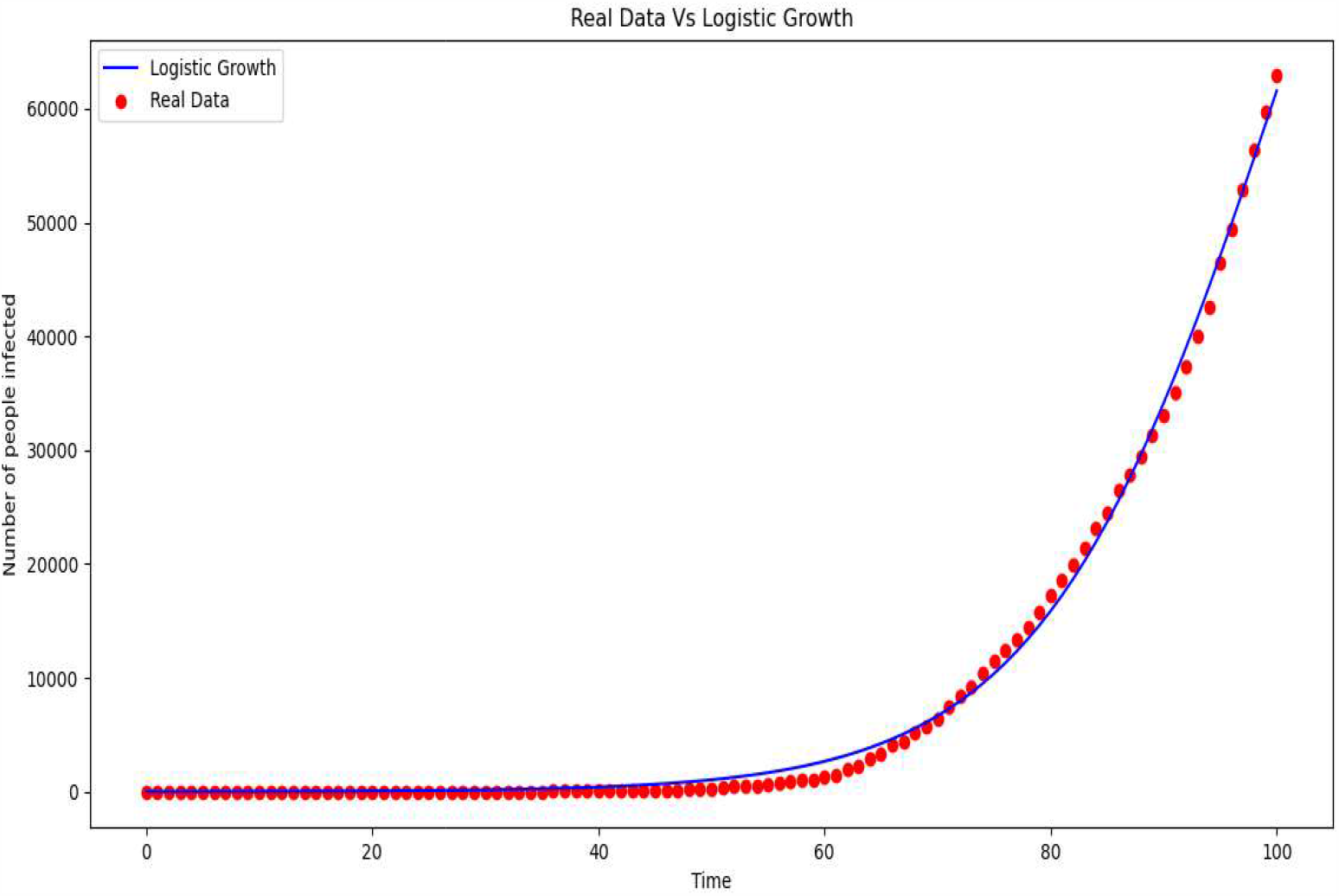
Logistic growth curve of COVID-19 in India

**Figure 3:**
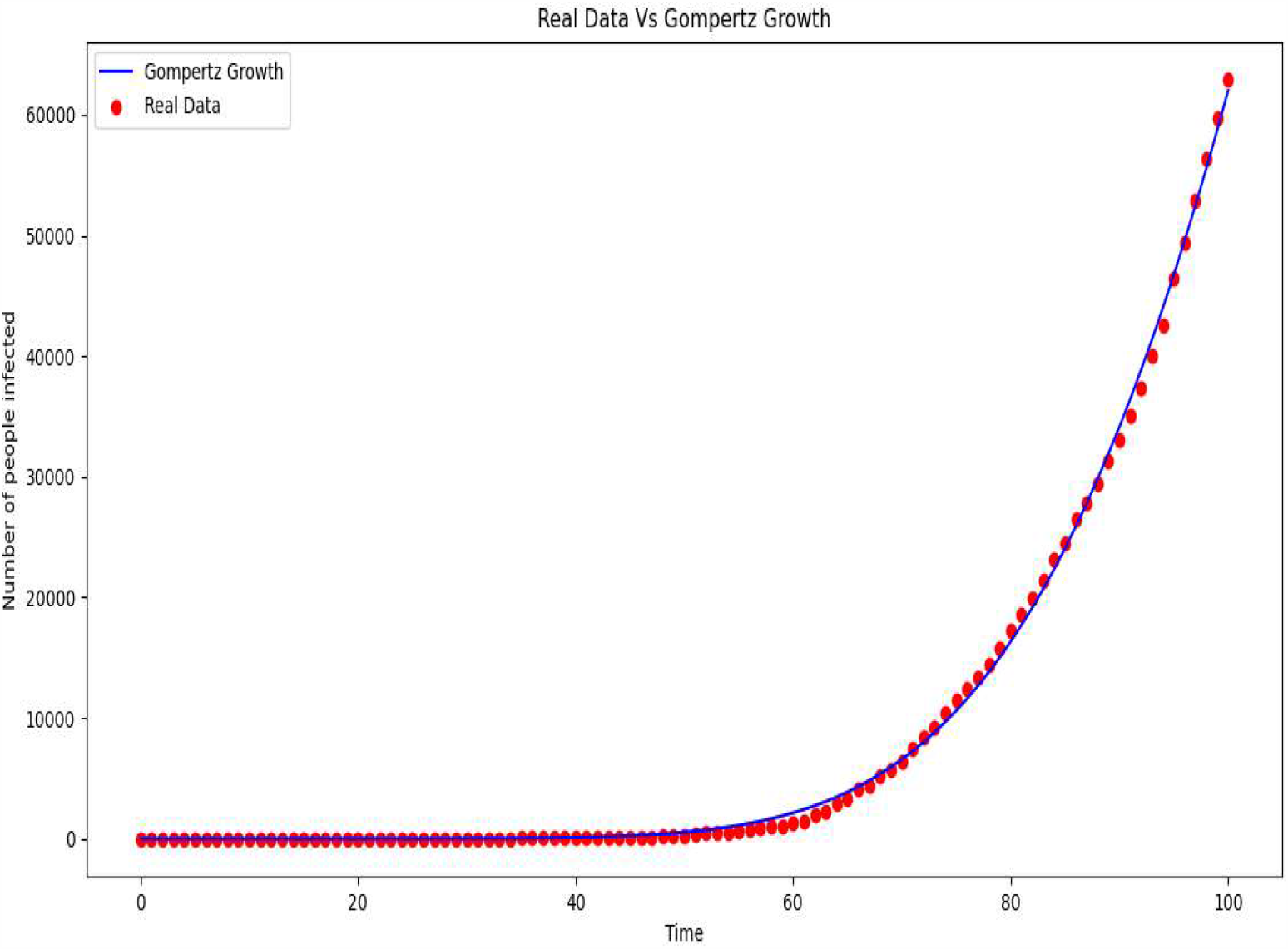
Gompertz growth curve of COVID-19 in India

**Figure 4:**
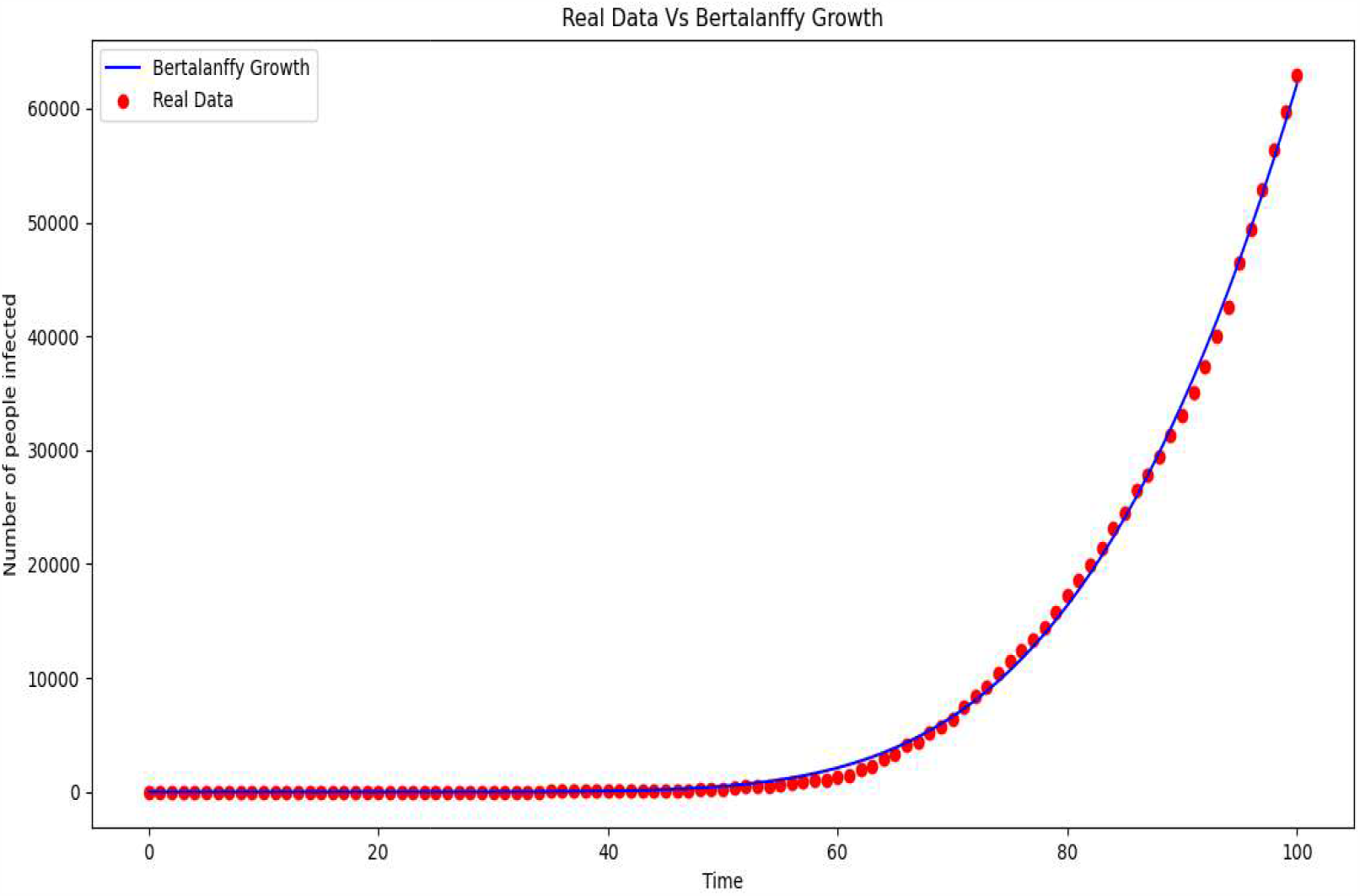
Bertalanffy growth curve of COVID-19 in India

Where ***I***_***0***_ is the initial number of cases, ***k*** is the carrying capacity of an epidemic, ***r*** is the growth rate.

As of 21 May 2020, India’s Case fatality rate is 5.3989

Quartiles are calculated. Then the Minimum = 0.00, Lower quartile (25%) = 0.7878 (Q1), Median (50%) = 1.5918, Upper quartile 3 (75%) = 2.6569(Q3), Maximum = 8.4430.

## IV. DISCUSSION AND RECOMMENDATIONS

Logistic model (R^2^=0.9976), Gompertz model (R^2^=0.9989) and Bertalanffy model (R^2^=0.9990) can better predict the development trend of COVID-19 in India in the later stages of the epidemic than that of Exponential model (R^2^ = 0.1605). Based on the values of coefficient of determination, the best model fitted is Bertalanffy growth model. As per the best model, the impact of the virus will be greater than the population of Bahrain, lasting for more than a year, before it leaves India for good.

As per Bertalanffy growth model the expected number of cases at infection end is 16,09,200 and infection end is predicted around July 2023. Number of infections on 20 June 2020 is predicted as 2,91,847. These predictions hold only under the scenario of lockdown, where controlling measures for the pandemic situation of COVID-19 are solid and well founded. These predictions will probably change due to extrinsic effects like new lockdown policies including lockdown lifting, virus mutation, vaccine discovery and so on. From Case fatality rate based classification we can identify the regions under high risk. We recommend that degrees of relaxation in lockdown restrictions could be allotted to each region based upon the group they belong to.

## V. CONCLUSION

Growth model forecasting points at the steady growth of COVID-19 in India. The rate of spread would have decreased in the scenario of lockdown. If the current trend continues, COVID-19 is going to hit lakhs in India. Larger values of CFR in some states can be a result of the scanty capacity of the surveillance system to set off a timely response. We all should get to work to spread awareness for preventing the spread of coronavirus, to have a better future.

## Data Availability

https://www.kaggle.com/sudalairajkumar/covid19-in-india?select=covid_19_india.csv
https://ourworldindata.org/coronavirus

https://ourworldindata.org/coronavirus

## CONFLICT OF INTEREST

The authors declare that they have no known competing financial interests or personal relationships which might seem to have influenced the work reported in this paper.

## ACKNOWLEDGMENTS

We are grateful to Dr. K.K Jose, HOD Department of Biostatistics, St. Thomas College, Palai for the valuable comments and suggestions leading to improvement of this paper.

## Notes

### Competing Interest Statement

The authors have declared no competing interest.

### Author Declarations

St.Thomas College, Palai

